# Operationalizing Occupational Fatigue in Pharmacists: An Exploratory Factor Analysis

**DOI:** 10.1101/19008169

**Authors:** Taylor Watterson, Kevin Look, Linsey Steege, Michelle Chui

**Affiliations:** University of Wisconsin-Madison School of Pharmacy, Madison, WI, USA; University of Wisconsin-Madison School of Nursing, Madison, WI, USA

## Abstract

**Introduction:** The Quadruple Aim recognizes that caring for the healthcare employee is necessary to optimize patient outcomes and health system performance. Although previous research has assessed pharmacists’ workload, this study is the first to describe pharmacist occupational fatigue – a characteristic of excessive workload that inhibits workers’ abilities to function at normal capacity. The purpose of this study was to describe occupational fatigue in pharmacists using exploratory factor analysis (EFA) – assessing whether dimensional structures used to describe occupational fatigue in other health professions fit pharmacist perceptions.

**Methods:** A model was created to conceptualize ‘fatigue’ domains found in the literature. A priori, the two domains identified were physical fatigue (ex. physical discomfort), and mental fatigue (ex. trouble thinking clearly). These domains were operationalized and used to create a paper survey that was distributed to licensed pharmacists at a pharmacy conference. An EFA was conducted to identify the key domains underlying pharmacist perceptions of fatigue.

**Results:** A total of 283 surveys were distributed and 115 were returned and usable. Respondents were primarily white, female, worked 9.52 hours-per-day on average, and mean age of 39-years-old. The EFA suggested a statistically significant two-factor model (χ^2^ 9.73, p=0.28; TLI 0.998, RMSEA 0.048), which included physical fatigue (α = 0.87) and mental fatigue (α = 0.82).

**Discussion:** The EFA yielded a structure similar to what was anticipated from the literature with physical and mental fatigue dimensions. This is just the first step in promoting systematic interventions to prevent or cope with fatigue and prevent the downstream patient, pharmacist, and institutional outcomes. By addressing fatigue and caring for employees, health care systems can take steps to work toward the Quadruple Aim.

## INTRODUCTION

In 2008, colleagues at the Institute for Healthcare Improvement promoted a collection of health care initiatives entitled the Triple Aim, which encompassed: “improving the individual experience of care, improving the health of populations, and reducing the per capita costs of care for populations”.^1^ These goals were promoted as a way to integrate and innovate the United States healthcare system but have been criticized for not addressing a key component of the health delivery system—the healthcare employee.^2,3^ In 2014, a fourth aim was proposed to establish a Quadruple Aim that emphasizes the well-being of the healthcare professional. The Quadruple Aim recognizes that caring for the healthcare employee is a prerequisite to optimizing patient outcomes and health system performance—attributing excessive expectations of health care professionals to increased burnout and negative health outcomes for providers and patients.

One negative outcome that is important to address is occupational fatigue—described as “a multidimensional state that arises in workers who are exposed to excessive demands through their work tasks, environment, and schedules, and that can interfere with workers’ physical and cognitive abilities and their ability to function at their normal capacity.”^4^ Occupational fatigue occurs across a continuum, ranging from acute fatigue to chronic fatigue, and is multidimensional, including mental, emotional, and physical fatigue.^5^

Fatigue is crucial to assess as evidence suggests that fatigue is associated with decreased safety in the workplace.^6^ In their seminal report, “To Err is Human: Building a Safety Health System,” the National Academy of Sciences highlighted the importance of safe environments for both healthcare workers and patients and noted that fatigue may be a contributing factor to hazards or injuries for both healthcare workers, but also to the patients they care for due to its association with decreased performance.

Originating outside of healthcare in transportation literature including driving and aviation, fatigue has been long associated with increased risks to safety.^7^ In nursing, fatigue caused by long shifts exceeding twelve hours, was associated with a significantly increased risk of making an error.^8^ Similarly, research has shown that fatigued physicians: have delayed response times and diagnostic ability, are more likely to sustain needle stick injuries, are more likely to make errors, and are more likely to experience road traffic incidents at the end of their shifts.^9^ A study of nursing leadership found that nurse leader fatigue impacted decision-making, work-life balance, and turnover intent.^10^

To date, no research exists evaluating occupational fatigue in pharmacists, yet pharmacists face unique challenges compared to other healthcare professionals.^11–19^ In addition to dispensing medications, pharmacists in community settings often make over-the-counter product recommendations, provide vaccinations, ensure safe and adequate dosing of medications, check for interactions, and provide comprehensive medication reviews for patients to ensure cohesive and holistic medication management. However, pharmacists are expected to fulfill these requirements while being in separate geographic locations from prescribers and other healthcare providers, with limited or no access to patients’ electronic medical records. Additionally, pharmacists cannot control their rate of work— patients do not make appointments to fill or pick up prescriptions and many locations have numerous access points for patients to interact with the pharmacy staff (several telephone lines, drop off windows, pick up windows, drive through lanes, fax, electronic communication, etc.). Finally, most community pharmacies are for-profit organizations, which makes them a unique healthcare setting and places additional stressors on the pharmacists with regards to competition, labor, and efficiency. Although previous research has assessed these and other components of pharmacists’ workload, this study is the first to describe occupational fatigue.^12–15^

Given the push from the Quadruple Aim and the potential implications for patient and employee safety in other professions, there is a timely need to assess pharmacist fatigue and create tools to address this growing problem. The purpose of this study was to describe occupational fatigue in pharmacists using exploratory factor analysis—assessing whether dimensional structures used to describe occupational fatigue in other health professions fit pharmacist perceptions.

## METHODS

### Study Design

This study combined the ideas of the Systems Engineering Initiative for Patient Safety (SEIPS) model, Swedish Occupational Fatigue Inventory (SOFI), and fatigue literature from other healthcare professions to create a conceptual model of occupational fatigue (Figure 1).^20–23^ The SEIPS model illustrated the impact of excessive demands work on pharmacist work and processes. This then yields possible implications for both pharmacist and patient outcomes.^24^ The SOFI scale is commonly used in fatigue literature had has been used in numerous applications.^25–28^ The SOFI lists items of fatigue which include feeling: overworked, drained, spent, worn out, breathing heavily, out of breath, sweaty, experiencing palpitations, aching, having stiff joints, numbness, tense muscles, uninterested, indifferent, passive, lack of concern, sleepy, yawning, drowsy, and falling asleep.^20^ A comprehensive literature review was conducted to find other major themes related to fatigue in healthcare professionals to explore during a developmental interview process.

**Figure 1.**
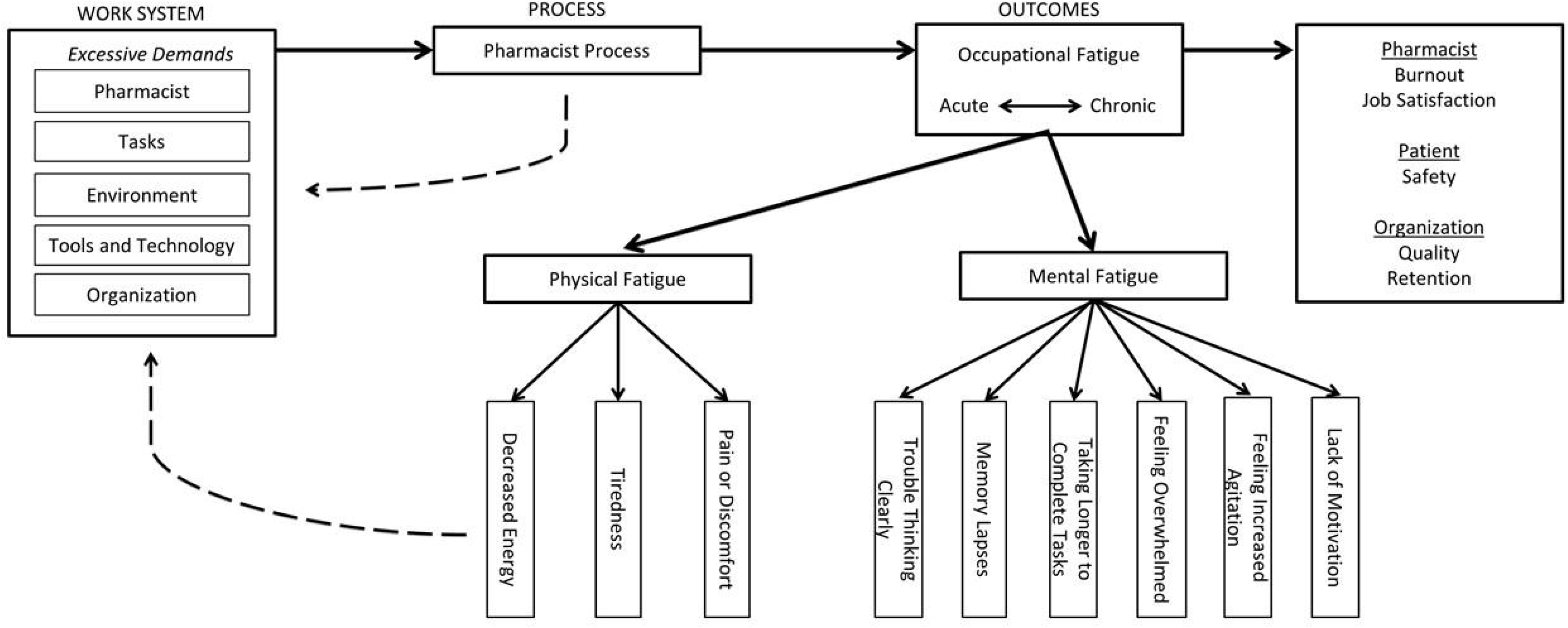
Conceptual Model

To understand how the concepts of occupational fatigue and items of the SOFI were perceived by the target population, developmental interviews were conducted with two community pharmacists.^29–40^ Respondents were asked to describe changes to their mental and physical states that occurred over the course of the day. This helped to translate the ideas of the conceptual models into vocabulary and terms that would be understood by respondents. Terms and vocabulary that affiliated with fatigue inventory items from the SOFI were used to generate survey questions. Although the wording and terminology were organically created using developmental interviews, the questions and model were still founded in the concept of “occupational fatigue” as previously defined. The instrument was pre-tested seven times with a convenience sample of pharmacist colleagues. Two cognitive interviews were conducted with pharmacists to understand how respondents interpreted survey questions. The respondents were asked to “think aloud” as they were using the instrument and often probed with additional questions.^35^ Respondent feedback on confusing wording, vague terminology, and general assumptions provided valuable feedback that was incorporated into the final instrument. This study was approved by the University of Wisconsin-Madison’s Institutional Review Board (IRB) prior to data collection.

### Setting and Participants

The survey was administered in April of 2018 at a two-day pharmacy education conference hosted by a state pharmacy association. A paper survey was distributed to be completed by hand and individuals were given a small, non-contingent incentive, that they were able to keep regardless of completion of the survey. A total of 283 surveys were distributed.

### Statistical Methods

The paper surveys were entered into the electronic database by a single individual (TLW). Data entry verification was conducted by verifying 20% of the total responses. The data analyses were completed using R version 3.4.3. (The R Foundation for Statistical Computing Platform). Principal Components Analysis (PCA) and Exploratory Factor Analysis (EFA) were conducted to understand the data’s underlying structure that was underpinning pharmacist responses to the survey and generate a conceptual model. PCA is a variance maximizing mathematical procedure that provided insight into the number of principal components that accounted for a majority of the variance in the data and informed the number of factors to test in an EFA. The EFA was conducted to assess the underlying structure for pharmacist fatigue while accounting for survey measurement error (i.e. the idea that the survey scales may not be perfectly reliable). The EFA analysis utilized Maximum Likelihood (ML) estimation and promax oblique rotation.

An EFA factor loading of 0.4 was chosen as a conservative cutoff and items with a factor loading less than 0.4 were removed from the dataset and the analysis run again.^41^ The survey went through three iterations of item reduction before reaching acceptable factor loadings (all items greater than 0.4), goodness-of-fit statistics (significant chi-square and p-value > 0.05, TLI > 95%, RMSEA < 0.05), and internal consistency of items within the domains (Chronbach’s Alpha > 0.80).^42^

## RESULTS

### Participants

Of the 283 surveys distributed, 115 were returned and used in the analysis—accounting for a 40.6% response rate. Respondent characteristics are summarized in Table 1. Respondents were primarily white (89%), female (60%), and 39-years-old on average. Respondents worked 9.52 hours-per-day on average and half (50%) worked in a hospital or institutional setting. A chi-square goodness of fit test indicated a significant difference in the distribution of practice settings (X^2^ 77.13, df = 4, p < 0.0001), which was notably greater for the hospital/institution setting.

**Table 1.** Respondent Characteristics

|  | Respondents |  |
| --- | --- | --- |
|  | N = 115 | Percentage (%) |
| **Practice Setting** |  |  |
| Community Pharmacy | 13 | 11.2 |
| Long Term Care | 4 | 3.5 |
| Ambulatory/Outpatient | 29 | 25.2 |
| Hospital/Institution | 57 | 49.6 |
| Other | 12 | 10.4 |
| Specialty, Health System, PBM, Industry, Academia, Government, Health Plan, Hospital Administration, Home Infusion |  |  |
| **Ethnicity** |  |  |
| Asian/Pacific Islander | 9 | 7.7 |
| Black | 1 | 0.9 |
| Hispanic | 3 | 2.6 |
| White/Non-Hispanic | 104 | 88.8 |
| **Gender** |  |  |
| Male | 46 | 40 |
| Female | 69 | 60 |
| **Scheduled Breaks in Work Day** |  |  |
| Yes | 81 | 70.4 |
| No | 34 | 29.6 |

### Principal Components Analysis

The standard deviation, eigenvalue, proportion of total variance, and cumulative proportion of variance are listed in Table 2. The scree plot and eigenvalues of the complete 12-item data set suggested four principal components that accounted for 73.18% of the total variance. The suggested four principal components of the 12-item data influenced the EFA procedure.

**Table 2.** Principal Components Analysis of Original 12-Item Instrument

|  | Standard Deviation | Eigenvalue | Proportion of Variance | Cumulative Proportion of Variance |
| --- | --- | --- | --- | --- |
| Component 1 | 2.605 | 6.788 | 0.450 | 0.450 |
| Component 2 | 1.358 | 1.845 | 0.122 | 0.573 |
| Component 3 | 1.1233 | 1.262 | 0.0837 | 0.656 |
| Component 4 | 1.066 | 1.135 | 0.0753 | 0.732 |
| Component 5 | 0.965 | 0.932 | 0.0618 | 0.794 |
| Component 6 | 0.837 | 0.701 | 0.046 | 0.840 |
| Component 7 | 0.813 | 0.662 | 0.044 | 0.884 |
| Component 8 | 0.708 | 0.501 | 0.033 | 0.917 |
| Component 9 | 0.620 | 0.384 | 0.025 | 0.943 |
| Component 10 | 0.569 | 0.324 | 0.022 | 0.964 |
| Component 11 | 0.543 | 0.295 | 0.020 | 0.984 |
| Component 12 | 0.495 | 0.245 | 0.016 | 1.000 |

### Exploratory Factor Analysis

The factor loadings for a 2-factor structure are presented in Tables 3 and 4 with the factor associations highlighted. Three rounds of EFA and item reduction were conducted, removing a total of 5 items from the original 12. Item reduction concluded when the structure fit-statistics indicated a statistically significant model fit.

**Table 3.** Maximum Likelihood Estimation for 2-Factor Structure of 12-Items

| Item # | Item | EFA Constructs |  |
| --- | --- | --- | --- |
|  |  | ML1 | ML2 |
| Q14_54 | Feel that your energy decreased over the course of the day? | 0.99 | -0.14 |
| Q14_55 | Feel more tired later in the day than you did at the beginning of the day? | 1.00 | -0.24 |
| Q14_56 | Feel pain or discomfort anywhere in your body? * | 0.41 | 0.18 |
| Q14_65 | Have times where you felt generally fatigued? | 0.65 | 0.19 |
| Q14_57 | Have trouble thinking clearly at work, even for a short time? | 0.04 | 0.76 |
| Q14_58 | Have times where you forgot whether or not you had completed a task? | -0.20 | 0.87 |
| Q14_59 | Have times where you spent longer completing a task later in the day than you would at the beginning of the day? | 0.12 | 0.51 |
| Q14_60 | Have times where you felt that you could not keep up with your work? * | 0.10 | 0.49 |
| Q14_61 | Have times where you felt more impatient later in the day than you did at the beginning of the day? * | 0.23 | 0.37 |
| Q14_62 | Have times where you felt that you were not performing at your best? | 0.17 | 0.65 |
| Q14_63 | Find it necessary to take short-cuts when providing patient care? * | -0.22 | 0.74 |
| Q14_64 | Have times where you felt that you were not able to go above and beyond standard patient care? * | 0.12 | 0.55 |
| SS Loadings |  | 2.72 | 3.32 |
| Proportion of Variance |  | 0.23 | 0.28 |
| * Indicates item dropped following exploratory factor analysis |  |  |  |

**Table 4.** Pharmacist Occupational Fatigue Survey Items Retained in 2-Factor Structure

| Item # | Item | EFA Constructs |  |
| --- | --- | --- | --- |
|  |  | ML1 | ML2 |
| Q14_54 | Feel that your energy decreased over the course of the day? | 0.94 | -0.05 |
| Q14_55 | Feel more tired later in the day than you did at the beginning of the day? | 0.93 | -0.13 |
| Q14_65 | Have times where you felt generally fatigued? | 0.59 | 0.29 |
| Q14_57 | Have trouble thinking clearly at work, even for a short time? | -0.04 | 0.90 |
| Q14_58 | Have times where you forgot whether or not you had completed a task? | -0.19 | 0.84 |
| Q14_59 | Have times where you spent longer completing a task later in the day than you would at the beginning of the day? | 0.08 | 0.56 |
| Q14_62 | Have times where you felt that you were not performing at your best? | 0.19 | 0.61 |
|  | SS Loadings | 2.16 | 2.30 |
|  | Proportion of Variance | 0.31 | 0.33 |
|  | Cronbach Alpha | 0.87 | 0.822 |
|  | Chi Square | 9.73 |  |
|  | P-Value | 0.28 |  |
|  | TLI | 0.998 |  |
|  | RMSEA | 0.048 |  |
|  | BIC | -28.23 |  |

The two-factor model yielded a simple structure with all items loading on only one factor or the other (loadings are all greater than 0.50) with low cross-loadings (no secondary loadings greater than 0.20) (Table 3). The structure presented a factor correlation of 0.58. According to the likelihood chi square and associated p-value (X^2^ 9.73, p= 0.28), the two-factor model fits the data and fails to reject the null hypothesis that there is no difference between the data and the model. When comparing the model to “baseline,” the TLI indicated that the two-factor model fit better than no model at all (99.8%). When considering approximate fit, the RMSEA suggested that the two-factor model fit the data well (0.048). The Cronbach’s Alpha for the two factors indicated good item correlation within each factor (0.87 and 0.82 respectively). The final 2-factor model is visually depicted in Figure 2.

**Figure 2.**
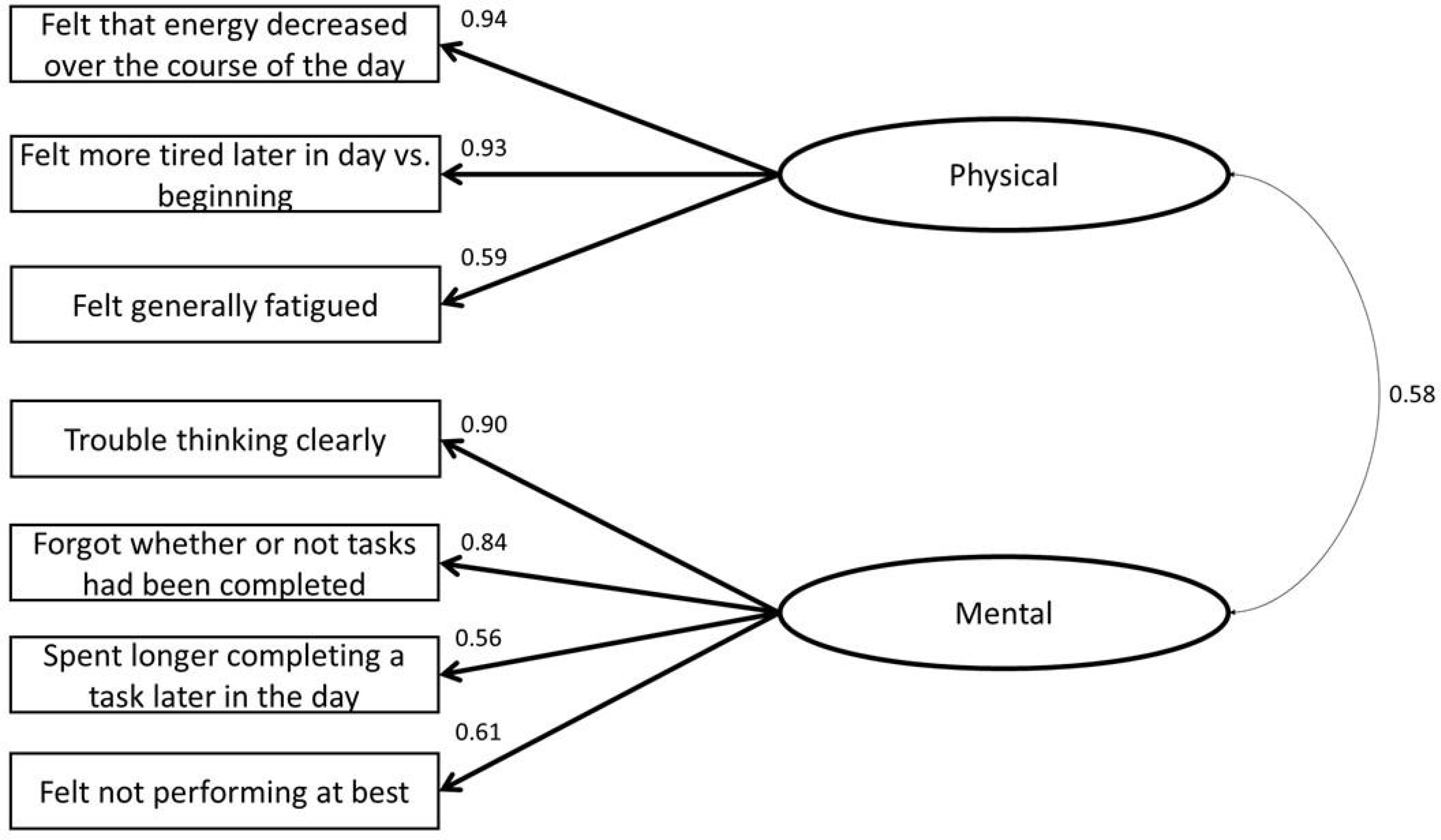
Maximum Likelihood Two Factor Model

The survey results and exploratory factor analysis provided insight into the structure underpinning pharmacist fatigue. As depicted in Figure 2, the factors in the two-factor structure were named “physical” and “mental.” The “physical” factor contained the items: felt that energy decreased over the course of the day, felt more tired later in the day versus the beginning of the day, and felt generally fatigued. The “mental” factor contained the items: trouble thinking clearly, forgot whether or not had completed a task, spent longer completing a task later in the day, and felt not performing at his/her best.

## DISCUSSION

This study adds to the occupational fatigue literature, discussing occupational fatigue particularly in healthcare professionals. The EFA presented a two-factor model of best fit—characterizing the physical and mental dimensions of occupational fatigue that was similar to the a priori conceptual model and nurse fatigue literature with physical and mental fatigue dimensions. ^4–8, 21, 43^

During item reduction, the one item addressing pain or discomfort surrounding fatigue was removed for its lack of strong factor loading. The assessment of the pain/discomfort suggests that this “physical” concept may not be as pervasive in pharmacists as originally anticipated. This is interesting given that, often, pharmacists stand for the majority of their shifts and look at computer screens for prolonged periods of time—tasks and physical factors of the work environment that may be associated with pain. The results of this study were in agreement with the literature surrounding nurse fatigue, which highlighted that nurses reported higher levels of mental fatigue rather than physical fatigue.^43^ It’s interesting to note; however, that nurse and pharmacist tasks and workload differ greatly. While nurse workload may require the employee to lift a patient in and out of bed, establish IV lines, and manage patient comfort, the pharmacist utilizes cognitive functions to assess dosing regimens, compute doses, check for reactions, and provide consultations. The range of tasks required is different for the two professions and it was expected that this would be differentiated in the fatigue rating.

Other items removed from the a priori model mapped to the mental fatigue dimension and, in general, suggested statements of a more “sensitive” nature—for example, admitting to taking short cuts when providing patient care may make the pharmacists’ liable in the event of error. Assuming the respondents’ answers were accurate and free from bias, the results suggest that pharmacists may not perceive their fatigue to impact their provision of patient care. Cognitive interviews suggested that pharmacists would not purposefully provide inadequate patient care services as a result of fatigue, but that fatigue may cause a lapse in judgement or ability. Some interview participants indicated that excessive workload demands, feeling “foggy,” and tired, may cause them to not go “above and beyond” standard patient care as they would normally like. For example, one pharmacist anecdotally mentioned during a developmental interview that near the end of a 12-hour shift, she was so “drained” and had been “working at maximum capacity” for so long that her patient counseling sessions were minimal and did not cover the dearth of information that she might have earlier in the day.

Pharmacist well-being is a fairly novel concept that has been gaining traction, especially with discussions of burnout, resiliency, and job satisfaction. Yet in the moment, pharmacists may not be aware of fatigue related short-cuts or lapses in judgement that pose risks to patient safety—only identifying these vulnerabilities upon later reflection. The increased risk for error found in other fatigued healthcare professionals presents a similarly alarming risk to patient safety in fatigue pharmacists who are not even aware of their state.^8,9^

Beyond the patient, other considerations include the pharmacist outcomes possible as a result of occupational fatigue. For example, fatigue may have implications for burnout and turnover intent, as well as risks to safety in the form of needle sticks or road traffic incidents.^9,10^ Future discussions need to consider liability for fatigue related incidents—whether the individual or the organization is responsible for negative outcomes. Organizations may also be interested in assessing employee fatigue for other outcomes including: performance indicators, absence, and job turnover.

Overall, pharmacist occupational fatigue presents alarming and emergent risk to patient safety, employee safety, and public health. This study supports the tenants of the Quadruple Aim by focusing on the wellbeing of the health care practitioner, the pharmacist.^3^ The study’s conceptual model supports the notion that pharmacist occupational fatigue acts as a proximal outcome that occurs prior to other, more distal, outcomes—suggesting that assessing pharmacist fatigue may be a necessary prerequisite prior to enhancing the patient experience, the increasing the health of the population, and reducing costs.

### Limitations

It is important to consider the limitations to this research study when assessing the results, significance, and potential impact. When conducting an EFA, particularly ML estimation, there are assumptions that the sample expresses continuous and multivariate normality. Given that the survey utilized a Likert-scale, which is non-normal, but instead categorical, the goodness-of-fit indices may be inflated.

However, the Likert scale was based on a continuous and normally distributed variables, and was deemed usable for the ML estimation. One potential limitation of this study was the population that was sampled—pharmacists attending an educational conference. There may have been respondent bias— those who were inclined to participate in research or had strong feelings towards fatigue were more likely to complete the survey. The sample may lead to limitations of the generalizability of the survey results—describing fatigue in individuals that attend pharmacy conferences. The fatigue model may not be descriptive of experienced pharmacists or those who work in a setting which has more set/fixed scheduling (such as a retail community pharmacy).

### Future Research

Numerous opportunities exist for future research to validate the occupational fatigue measurement tool. Additional studies are needed to confirm the factor structure identified in this study with larger and more diverse samples of pharmacists. Objective measures that have been used in other industries and that assess physiologic markers (e.g. eye tracking) or performance changes (e.g. vigilance testing) associated with fatigue may also be valuable for monitoring and better understanding pharmacists’ experiences with fatigue and associated risks to safety.^44–46^ In addition, given that this study did demonstrate that pharmacists experience occupational fatigue (mental and physical), additional research is needed to assess the relationships between pharmacist fatigue levels and patient, pharmacist and organizational outcomes.

## CONCLUSION

This was the first study to assess occupational fatigue. A conceptual model was created to describe this concept and guided the development of a survey to assess the frequency and perceptions of occupational fatigue in pharmacists. Exploratory factor analysis identified two related dimensions: physical and mental fatigue and was similar to fatigue literature found in other healthcare professionals such as nurses. This is just the first step in promoting systematic interventions to prevent or cope with fatigue and prevent the downstream patient, pharmacist, and institutional outcomes. By addressing fatigue and caring for employees, health care systems can take steps to work toward the Quadruple Aim.

## Data Availability

Not applicable

